# Shortened Cortical Silent Period in Children with Attention Deficit Hyperactivity Disorder

**DOI:** 10.64898/2026.05.26.26354157

**Authors:** Darius S. Feier, Donald L. Gilbert, Deana Crocetti, Karlee Y. Migneault, David A. Huddleston, Paul S. Horn, Stewart H. Mostofsky, Steve W. Wu

## Abstract

**Background and Objective:** In ADHD, a heterogeneous neurodevelopmental condition, behavioral and motor manifestations may reflect multiple inefficient or perturbed inhibitory systems. To evaluate Transcranial Magnetic Stimulation (TMS) evoked cortical silent period (CSP) duration, an indicator of GABA_B_ receptor-mediated inhibition in motor cortex, as a potential biomarker of Attention-Deficit/Hyperactivity Disorder (ADHD) in children.

**Method:** We retrospectively analyzed TMS data, obtained using both round and figure-of-8 coils, from three cross-sectional studies conducted in 8- to 12-year-old children with ADHD (n=79; 10.7 +/− 1.5 years old) and age-and-sex-matched typically-developing controls (n=96; 10.5 +/− 1.4 years old).

**Results:** Median CSP was 32% shorter in ADHD (p=0.02). Regression analysis demonstrated a relationship between shorter CSP and both lower active motor thresholds (p < 0.0001) and more severe hyperactivity symptom rating (p = 0.026). Test-retest CSP measures in 83 children showed moderate reliability (intraclass correlation 0.77 [ADHD], 0.75 [controls]).

**Conclusion:** TMS-evoked CSP may be a useful biomarker in future investigations of ADHD subtypes, domains of impaired function, or treatment outcomes.

## INTRODUCTION

Attention-deficit/hyperactivity disorder (ADHD), characterized by symptoms of inattention, impulsivity, and hyperactivity, is the most common neurodevelopmental disorder, diagnosed in over 10% of children in the United States.^1^ To improve both biological understanding of neurodevelopmental disorders like ADHD and accelerate more targeted treatments, several working groups have promoted biomarker development.^2^ A critical obstacle to this goal is that ADHD is a heterogeneous, clinically-diagnosed condition. Its symptoms exist on a spectrum with typical development, and the diagnosis has high prevalence in a wide variety of genetic and acquired conditions.

Over the past two decades, multiple neurophysiological measures have been investigated as possible diagnostic or treatment biomarkers for ADHD,^3^ with the most promising two Transcranial Magnetic Stimulation (TMS) measures being resting short interval cortical inhibition (SICI)^4^ and task related up-modulation (TRUM) from resting to active states, particularly when having to inhibit a prepotent motor response.^5^ However, as with biomarkers derived using electroencephalography (EEG) or neuroimaging,^3^ these TMS measures show overlap between ADHD and TD controls. To capture and stratify diagnostic subgroups or predict treatment outcomes, additional TMS biomarkers may be useful.

One such biomarker candidate is the duration of the TMS-evoked cortical silent period (CSP), which can be easily measured in children by delivering a single TMS pulse over motor cortex (M1) while the child pre-activates a target muscle. This induces an inhibitory response that temporarily silences the ongoing EMG activity, for approximately 20-200 msec.^6^ While SICI reflects function of GABA_A_ receptors,^7^ ionotropic chloride (Cl^-^) channel receptors that mediate fast, phasic inhibition,^8^ CSP reflects function of GABA_B_ receptors,^9^ which are metabotropic and mediate slower inhibition via presynaptic calcium (Ca^2+^) channels and postsynaptic inward-rectifying potassium channels.^10^ SICI and CSP may therefore may provide information on complementary inhibitory processes.

CSP has been investigated across a range of disorders as a potential physiological biomarker. A meta-analysis of 14 studies involving 267 epilepsy patients and 234 healthy participants found, paradoxically, that individuals with idiopathic generalized epilepsy and partial epilepsy have prolonged CSP (greater inhibition) compared to controls.^11^ In addition, one study showed treating children with new-onset epilepsy with sodium valproate reduced (normalized) the CSP, supporting its potential as a treatment biomarker.^12^ In contrast, compared to healthy matched controls, persons with psychiatric disorders such as major depressive disorder and obsessive compulsive disorder were found to have shorter CSPs.^13,14^ For treatment resistant depressive patients, baseline CSP has been shown to predict clinical response to electroconvulsive therapy.^15^ Taken together, these findings support the notion that CSP may reflect transdiagnostic processes underpinning neuropsychiatric symptoms.

In children with ADHD, a meta-analysis of 8 studies involving 248 participants with ADHD and 278 controls found a statistically non-significant reduction in CSP.^16^ The authors suggested CSP might reflect some compensatory mechanisms, but did not have severity scores or other physiological measures within individuals to help evaluate this. In this study, we aimed to determine whether ADHD might be linked to CSP duration and, further, whether symptom severity or other physiological measures might also account for its variance. We therefore combined our CSP data across multiple studies for this new analysis. In addition, to better estimate reliability, we performed CSP in a subgroup of participants at two visits, separated by approximately 21 days, and we compared CSPs obtained with round and figure-of-8 TMS coils.

## METHOD

### Participants

This analysis combines published^17,5^ and unpublished data obtained through studies at two collaborating institutions, Cincinnati Children’s Hospital Medical Center and Kennedy Krieger Institute, from 2007 to 2024, mostly through a long-standing prospective collaborative effort, ensuring consistency and reliability of data collection across these two sites. These studies were approved by Institutional Review Boards of both organizations (including through a single-IRB reliance agreement since 2020). Patients and caregivers provided informed consent for the research and publication of the results.

Right-handed, typically developing (TD) children and children with ADHD aged 8 to 12 years were enrolled. Diagnostic criteria were standard, rigorous, systematic, and consistent across sites, as described elsewhere.^17^ Inclusion and exclusion criteria were the same throughout this time period and across sites. ADHD diagnosis was confirmed using a clinician-administered structured diagnostic interview; with the DICA used from 2007 to 2012 and the K-SADS used from 2012 to 2024.^18,19^ Children with ADHD with presence of other psychiatric diagnoses, based on structured diagnostic interview were excluded, with exceptions for oppositional defiant disorder (ODD) and simple phobia, as were children with reading disabilities or full-scale IQ < 80. TD control children could have no neurological, psychiatric, or developmental diagnoses. Participants could have no other significant medical diagnoses. ADHD symptoms were assessed with the parent Conners, 3^rd^ edition.^20^ Board certified child neurologists (DLG, SHM) reviewed all clinical data to confirm diagnostic eligibility.

Children with ADHD taking stimulants were included, so as to ensure a more representative sample; those taking other psychoactive medications were excluded. Parents were asked to withhold stimulant medication the day prior to and the day of testing. Standard TMS contraindications (e.g., seizure, hearing impairment, implanted medical device, serious medical conditions) were also applied.^21^

### TMS Procedures

Study personnel at both sites trained together for consistency of motor physiology assessments. TMS was performed using Magstim 200 stimulators (Magstim, Whitland, Wales, UK) with Bistim modules. The TMS hardware and procedures were identical across sites during each phase of the studies, with one exception. During the first study, we used a 70mm figure-8 coil^17^ while a 90mm circular coil was used subsequently. For the figure-8 coil, the coil was placed over left primary motor cortex, tangential to the skull with handle backward, at 45 degrees to the midline and its center near the optimal position and orientation for producing an MEP in the right first dorsal interosseous muscle (FDI). For the circular coil, TMS coil placement was flat at the vertex, with the handle pointing posteriorly towards the occiput and counterclockwise current in the coil. To obtain CSP, investigators first measured active motor threshold (AMT). Then, while participants contracted their right FDI, five TMS pulses over primary motor cortex were delivered at an intensity of 150%*AMT. An electromyogram was recorded with surface electrodes, amplified, and filtered (100/1,000 Hz) (Coulbourn Instruments, Allentown, PA) before being digitized at 2 kHz and stored for analysis using Signal software and a Micro1401 interface (Cambridge Electronic Design, Cambridge, UK).

The 5 tracings were rectified and averaged into a single tracing, with CSP onset and offset identified visually. All individual tracings were analyzed blinded to diagnosis. Starting in 2021, participants were invited to return for a 2^nd^ study visit (minimum of 10 days, mean 21 days between visits) to repeat CSP measurement to assess test-retest reliability.

### Statistical analysis

Analyses were performed using SAS® statistical software version 9.4 (SAS Institute Inc. Cary, NC). Based on testing showing CSP was not normally distributed, the Wilcoxon Mann Whitney test was used to compare 1) age and CSP durations between diagnostic groups and 2) CSP duration between figure-8 vs. circular coil groups. The t-test was used to compare RMT vs. AMT between ADHD and TD. The Chi square test was used to compare sex proportions for the diagnostic groups. In order to explore the relationship between symptom severity and CSP duration, a lasso (Least Absolute Shrinkage and Selection Operator) regression analysis^22^ was performed. Advantages of the lasso regression include avoidance of overfitting while avoiding collinearity. The dependent variable was CSP, while the independent variables were site, coil type (circular vs figure-8), RMT, AMT, Conners Inattention and Hyperactive-Impulsive t-scores. Age and sex were not included as independent variables because Conners t scores are already adjusted for these variables by design. Finally, we used the Shrout-Fleiss test^23^ to calculate intraclass correlation and Pearson correlation to examine test-retest reliability for CSP.

## RESULTS

### Participants

A total of 79 children with ADHD (10.7 ± 1.5 years old; 42M:37F) and 96 TD children (10.5 ± 1.4 years old; 55M:41F) participated (Table 1). Demographics, self-reported, were balanced across groups, with 69% Caucasian, 20% African American, 5% Asian, and 6% Biracial. The diagnostic groups did not differ by age (p=0.27), sex (p=0.58), RMT (p=0.42) or AMT (p=0.47). Eighty-three children (44 ADHD [10.0 ± 1.4 years; 25M:19F] and 39 TD [9.9 ± 1.2 years; 23M:16F]) returned for a second visit to re-measure CSP.

**Table 1.** Demographics and Clinical Data.

|  | ADHD<br>(n=79) | TD<br>(n=96) | p |
| --- | --- | --- | --- |
|  | n (%) | n (%) |  |
| Male n (%) | 42 (53%) | 55 (44%) | 0.58 |
|  | Mean (SD) | Mean (SD) |  |
| Age | 10.7 (1.5) | 10.5 (1.4) | 0.27 |
| RMT | 62.9 (13.0) | 64.5 (13.0) | 0.42 |
| AMT | 42.7 (10.2) | 43.8 (9.7) | 0.47 |
| Conners Inattentive t score | 73.6 (11.1) | 47.9 (8.3) | <0.0001 |
| Conners Hyperactive/Impulsive t score | 73.2 (14.3) | 47.8 (7.2) | <0.0001 |

### Cortical Silent Period

When compared to TD children (median 77.7 ms, IQR 42.9 – 122.5), CSP duration was shorter in children with ADHD (median 53.0 ms, IQR 33.7 – 98.6; p = 0.02; Figure 1). Lasso regression analysis showed that CSP was significantly associated with AMT (β = 1.45, p < 0.0001) and Conners Hyperactive-Impulsive t score (β = −0.44, p = 0.026). That is, shorter CSP duration is associated with lower AMT and worse ADHD. Although coil type was not a significant variable in the lasso regression analysis, univariate comparison with the Wilcoxon Mann Whitney test showed that CSP duration is longer when the circular TMS coil was used (p = 0.03).

**Figure 1.**
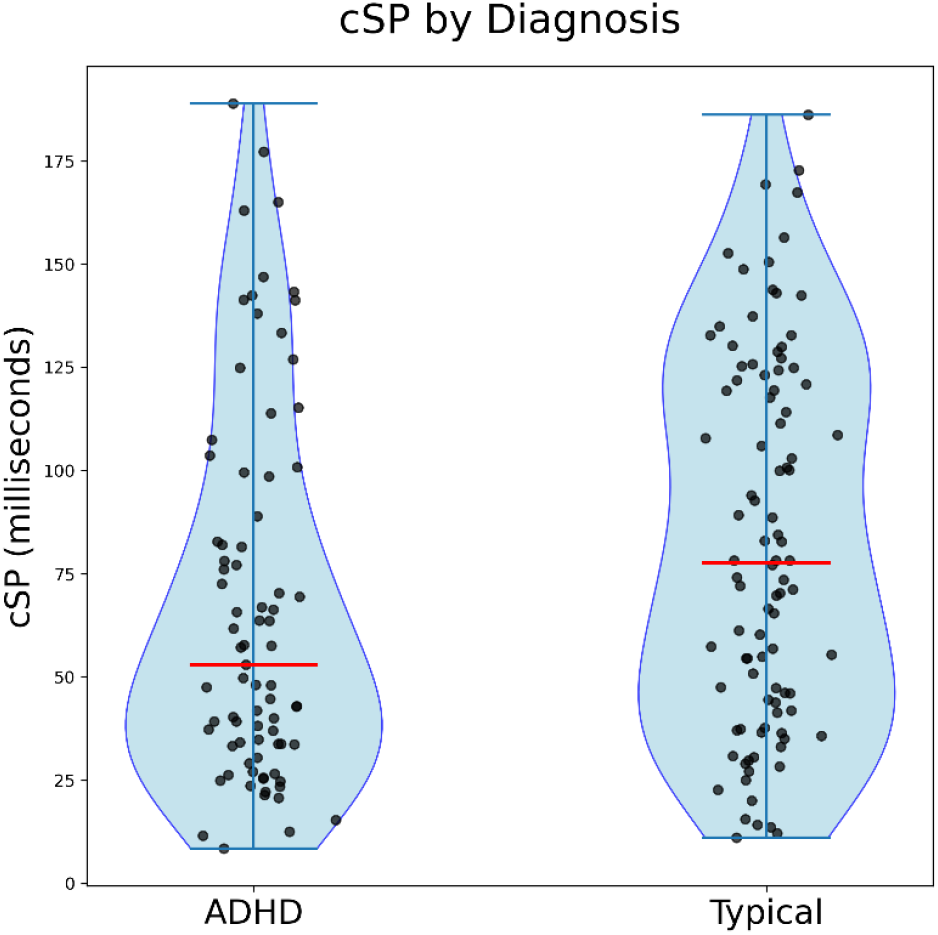
Cortical Silent Period. This violin plot shows the CSP by diagnosis group. The red line represents the median value. CSP duration is shorter in the ADHD group.

### Cortical Silent Period Reliability

CSP was found to have moderate test-retest reliability in repeated testing for both ADHD (Shrout-Fleiss ICC = 0.77 [95% CI: 0.61-0.87]; Pearson r = 0.77, p < 0.0001) and TD (Shrout-Fleiss ICC = 0.75 [95% CI: 0.57-0.86]; Pearson r = 0.76, p < 0.0001; Figure 2).

**Figure 2.**
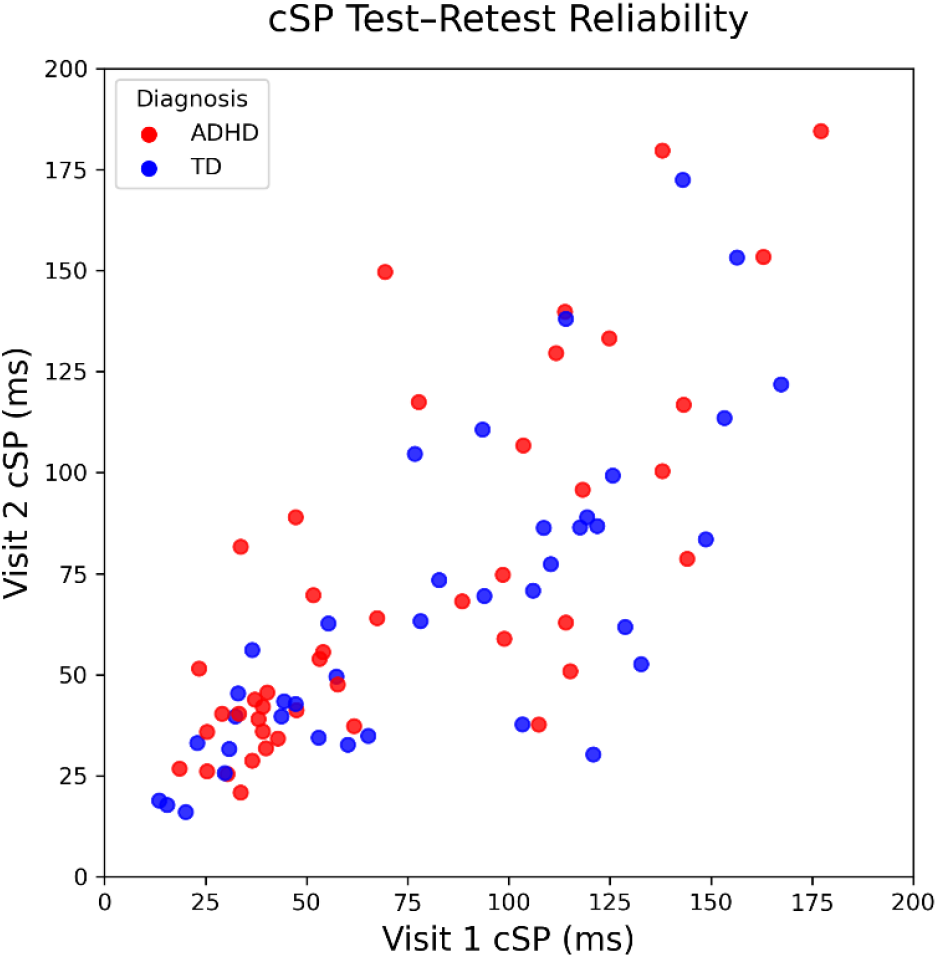
Cortical Silent Period (CSP) Test-Retest Reliability. Cortical Silent Period from two separate visits have moderate reliability for both ADHD (red) and TD (blue) children.

## DISCUSSION

This study shows a modest but statistically significant 32% reduction in the duration of the TMS-evoked CSP in 8- to 12-year-old children with ADHD compared to TD peers. We believe our results, obtained in two centers in two communities with demographics representative of the United States, generalize well to urban, school-age populations. Similar to other putative biomarkers,^2^ overlap between ADHD and controls is substantial (see Figure 1). Thus, no single CSP cut-off would show high sensitivity or specificity for diagnosis. However, the correlation with ADHD symptoms and the moderate test-retest reliability suggest that CSP could still be evaluated as a biomarker for treatment responses, or as part of a multimodal mechanistic study in isolated ADHD or in other diagnoses in which ADHD is prevalent.

### GABA and ADHD

The current study complements the existing knowledge of altered motor cortex physiology^24^ in children with ADHD. Motor development anomalies, particularly excessive overflow movements, long-observed in children with ADHD, have supported the idea of deficient or delayed development of inhibitory systems.^25^ Multiple prior studies have shown deficits in intracortical inhibitory activity in M1 SICI both at rest and during tasks.^16^ Reduced SICI correlates with higher ADHD symptoms and immature motor development.^17^ Imaging studies have also demonstrated altered relationships between motor cortex physiology and cortical GABA levels in ADHD.^26^ Herein we show, with group differences and symptom correlations with CSP, that aberrant inhibitory function in children with ADHD may also be reflected in impaired GABA_B_-mediated, slow presynaptic and postsynaptic inhibition. We also provide estimates for the first time in a large pediatric sample of the test-retest reliability of CSP, which is similar to reliability of SICI.^27^

### Factors that Influence CSP

The duration of CSP can be influenced by the stimulation intensity and by the direction of the TMS-induced current.^28^ In young children with high motor thresholds, due to TMS hardware limitations, it may not be possible to measure CSP using the commonly used 1.5*AMT protocol. Further research should evaluate the interaction between ADHD and TMS physiological measures such as CSP over a larger age-span and pulse intensity range.

A surprising finding in our study was the relationship between higher AMT and longer CSP. Within individuals, increasing the TMS stimulation intensity induces longer CSP duration. As is standard, to scale the CSP across individuals, we indexed pulse intensity to each individual’s measured AMT, the idea being that this accounts for cortical excitability during motor activation.^29^ Thus we did not expect to find this robust relationship. However, the other relevant factor is that in childhood RMT and AMT diminish with age.^30^ This is interpreted as a sign of cortical maturation – in more mature, fully myelinated cortex, MEPs are more readily recruited, and therefore a lower stimulation intensity is required for depolarization. Thus, within our cohort, “more mature” lower AMT is linked to both shorter CSP and, paradoxically, a higher (worse) ADHD rating scores.

### Limitations

In our extended collaboration studying young children with neurobehavioral and movement disorders, we have emphasized obtaining TMS measures using potentially scalable methods that are relatively inexpensive and easy to perform, especially in restless young children. As such, we did not utilize neuronavigation, nor did we track contraction intensity with visual EMG feedback. Although averaging a larger number of trials might have enhanced precision, we asked children during CSP to give us 5 maximum intensity squeezes, which we reasoned would be insufficient to fatigue most children. However, we cannot exclude a possibility that fatigue, or a startle in response to the TMS pulse, delayed the silent period offset. We also reasoned that, without visual monitoring, 5 maximum intensity muscle activations would be more consistent (less noisy) than 5 activations requested at some smaller fraction of maximum. Using a stronger intensity muscle contraction may have produced a modest difference compared to data generated by other researchers.

During the first data collection period, we used the figure-8 TMS coil, which is the most widely used configuration. Due to the hyperkinetic nature of ADHD children and difficulty maintaining a consistent angle and tangential position, we eventually transitioned to a circular TMS coil for easier coil placement. In our analysis, we found some evidence that the circular coil, which induces current more broadly across motor cortex, evokes a longer CSP. However, this finding on univariate analysis was not significant in the primary analysis regression, in which coil type did not account for the variance in CSP.

## Conclusion

We showed that GABA_B_-ergic CSP is abnormally shorter in ADHD children, supporting the notion that this population has deficient inhibitory activity. The inverse relationship between hyperactive symptom and CSP suggests that this biomarker could be helpful in tracking symptoms and potential intervention.

## Data Availability

All data produced in the present study are available upon reasonable request to the authors.

## FUNDING

This study was funded by National Institutes of Mental Health (R01MH095014; R01 MH095014; R01 MH078160). This manuscript is the result of funding in whole or in part by the National Institutes of Health (NIH). It is subject to the NIH Public Access Policy. Through acceptance of this federal funding, NIH has been given a right to make this manuscript publicly available in PubMed Central upon the Official Date of Publication, as defined by NIH.

## DATA SHARING AND ACCESSIBILITY

In accordance with NIMH and journal policies, all data are archived in the National Institute of Mental Health Data Archive.

## Notes

### Competing Interest Statement

The authors have declared no competing interest.

### Author Declarations

The Institutional Review Board of Cincinnati Children Hospital Medical Center and of the Kennedy Krieger Institute/Johns Hopkins Medical Institute gave ethical approval for this work.

